# The Regular Consumption of a Food Supplement Containing Miraculin Can Contribute to Reducing Biomarkers of Inflammation and Cachexia in Malnourished Patients with Cancer and Taste Disorders: The CLINMIR Pilot Study

**DOI:** 10.1101/2024.06.23.24309349

**Authors:** Ana Isabel Álvarez-Mercado, Bricia López Plaza, Julio Plaza-Diaz, Lucía Arcos Castellanos, Francisco Javier Ruiz-Ojeda, Marco Brandimonte-Hernández, Jaime Feliú-Batlle, Thomas Hummel, Samara Palma Milla, Ángel Gil

## Abstract

**Background:** Taste disorders are common in patients with cancer undergoing systemic therapy, persist during treatment and are associated with reduced food intake, increasing the risk of malnutrition. Cachectic syndrome, which is common in these patients and characterized by marked weight loss, anorexia, asthenia and anemia, is linked to the presence and growth of the tumor and leads to systemic inflammation. *Synsepalum dulcificum* is a plant whose berries contain miraculin, a glycoprotein that transforms sour tastes into sweet ones and could serve to ameliorate taste disorders in patients with cancer.

**Objective:** To evaluate the effect of the regular intake of Dried Miracle Berries (DMB), a novel food containing miraculin, on several biomarkers of inflammation and cachexia in malnourished patients with cancer and taste disorders receiving systemic antineoplastic therapy.

**Materials and methods:** Triple-blind, randomized, placebo-controlled clinical trial. Thirty-one patients with cancer of various etiologies receiving chemotherapy were enrolled in a pilot study and divided into three groups. The first group received a tablet containing 150 mg of DMB (standard dose); the high-dose group received a tablet of 300 mg of DMB, and the third group received a tablet with 300 mg of the placebo for three months before each main meal. Plasma levels of several molecules associated with inflammation and cancer cachexia were measured using the X-MAP Luminex multiplexing platform.

**Results:** The three groups showed a decrease in the plasma levels of IL-6, IL-1β, TNF-α, and PIF throughout the intervention, although the percentage change from baseline was greater in patients receiving a standard dose of DMB. In contrast, the CNTF concentration only decreased in the DMB standard-dose group. This group also presented the greatest reduction in the IL-6/ IL-10 ratio, while IL-15 and IL-10 increased in the groups treated with DMB but not in the placebo. Regardless of DMB consumption, sTNFR-II tended to decrease with treatment in patients who responsed well to the antineoplastic treatment. We did not find significant correlations between cytokines and sensory variables or dietary and nutritional status.

**Conclusions:** The regular consumption of a standard dose of the food supplement DMB containing miraculin along with a systemic antineoplastic treatment can contribute to reducing biomarkers of inflammation and cachexia in malnourished patients with cancer exhibiting taste disorders.

## 1 BACKGROUND

Cancer is one of the leading causes of death worldwide, accounting for almost 10 million deaths in 2020 (1 in 6 deaths) (1,2). The most common treatments for cancer are surgery, chemotherapy, and radiation. Other treatment options include targeted therapy, immunotherapy, laser therapy, or hormonal therapy (2). Chemotherapy and radiotherapy can cause taste and smell disorders by altering the structure of the pores of the palate with consequent thinning of the epithelium of the papilla (3), affecting taste nerves and salivary glands and destroying taste cells, including the eradication of proliferative progenitors (4). Those disorders alter the pleasure produced by taste and smell through the formation of conditioned aversions. In this regard, the term dysgeusia refers to quantitative or qualitative taste dysfunction, including taste distortions with bitter, metallic, salty, or unpleasant tastes (5,6). As a result, since food is perceived as unpleasant, a lower intake of food can contribute to weight loss and malnutrition in patients with cancer. Indeed, foods consumed can cause nausea, which negatively impacts the quality of life and nutritional status (5,7).

In severe cases, malnutrition can progress to cachexia, which is also a common condition in many patients with cancer (8); this complex metabolic disorder is characterized by a pronounced loss of muscle and fat mass, systemic inflammation, weakness and fatigue (9). Certain cancers also induce systemic reprogramming of the host’s energy metabolism. This leads to alterations in glucose, lipid and protein turnover. These metabolic imbalances are promoted by tumor-secreted and tumor-induced factors that promote cachexia (9).

Chronic systemic inflammation is also a critical component of cachexia. Inflammatory cytokines, particularly TNF-α, IL-6 and interferon-γ, are major drivers of many symptoms of the disease and are thought to be responsible for the metabolic changes associated with tissue loss in cancer wasting (10,11). In contrast, the cytokine IL-15, which is essential for the development, proliferation, and activation of immune cells, can also enhance the antitumor activity of immune cells and has been shown to possess significant antitumor potential (12). Interleukin-10 (IL-10), a multifunctional cytokine with multiple properties, has been extensively studied in various immunology and cancer biology fields. IL-10 is a pleiotropic cytokine that promotes cytotoxicity although high levels of IL-10 can inhibit antitumor responses (13). On the other hand, when present systemically, ciliary neurotrophic factor (CNTF) is involved in the induction of cachexia, induces the catabolism of stored fat, skeletal muscle protein, and liver glycogen, and decreases the circulating concentrations of several intermediary metabolites (5). Moreover, tumor-derived molecules such as proteolysis-inducing factor/dermcidin (PIF) have also been proposed as mediators of cancer cachexia. PIF induces protein degradation in skeletal muscle via the ubiquitin-proteasome proteolytic pathway (11). Notably, cancer cachexia is a strong independent cause of mortality in cancer patients (9) and those suffering from cancer cachexia are generally less tolerant to chemotherapies and radiotherapies, limiting their treatment options (14).

Unfortunately, cancer patients are involved in a vicious cycle in which illness provokes decreased food intake, malabsorption and/or increased loss of nutrients, leading to an increase in susceptibility to chemotherapy-induced toxicity. Ultimately, these complications, together with reduced mobility, fatigue, poor response to therapy and other complications, can further compromise nutritional status and the cachectic state becomes self-perpetuating (15).

Overall, products developed to prevent or alleviate cachexia should meet both the nutritional and sensory needs of cancer patients, promote the enjoyment of food, counteract malnutrition, stabilize weight and improve quality of life (7). In this sense, *Synsepalum dulcificum* (Daniell) is a plant whose berries contain miraculin, a glycoprotein that transforms sour flavors into sweet ones and limits bitter and metallic off-flavors, making food more palatable. In December 2021, the European Commission authorized the dried fruits of *S. dulcificum* (also known as DMB), which naturally contain the taste-modifying protein miraculin as a *novel food* (16). Recently, our group reported that regular consumption of DMB before each main meal for three months improved taste acuity and salty taste, leading to greater dietary intake (energy intake, fat quantity and quality), and ameliorated nutritional status (fat-free mass) and erythrocyte polyunsaturated fatty acid (PUFA) status (16). Indeed, an improvement in nutritional status could have a positive impact on inflammatory and cachexia states. In the present work, we aimed to evaluate the effect of regular consumption of DMB on several biomarkers of inflammation and cachexia in malnourished patients with cancer suffering from taste disorders and receiving systemic antineoplastic therapy.

## 2 METHODS

### 2.1 Study Design and Patients

The pilot CLINMIR study was designed to evaluate the efficacy and safety of the DMB food supplement on sensory function, nutritional status, dietary intake, quality of life and the fatty acid profile of erythrocytes in adult malnourished cancer patients with taste disorders undergoing active antineoplastic therapy (17). The complete protocol as well as the results of the major characteristics of patients and main outcomes, as well as the DMB composition, have been reported elsewhere (16,17).

The clinical trial protocol was approved by the Scientific Research and Ethics Committee of the Hospital University La Paz (HULP), Madrid (Spain) in version 1 in June 2022 and protocolled by the HULP Code 6164. This clinical trial was registered at http://clinicaltrials.gov (Clinical Trial NCT05486260). Briefly, a triple-blind, randomized, placebo-controlled intervention clinical trial with three arms was conducted. Thirty-one malnourished patients with cancer of various etiologies receiving systemic therapy and presenting taste disorders were included. Malnutrition diagnosis was assessed using the Global Leadership Initiative on Malnutrition (GLIM) criteria and morphofunctional assessment of disease-related malnutrition (18,19). Sensory disturbances were assessed by electrogustometry (20). The inclusion criteria included adult patients with cancer and systemic antineoplastic treatment for at least three months who had a weight loss ≥ 5%. All of them were capable of oral intake of food and drinks. The exclusion criteria were patients with cancer participating in another clinical trial, with enteral or parenteral nutrition, major gastrointestinal, metabolic, neurological and mental diseases, eating disorders, and severe digestive toxicity due to treatment with chemo-radiotherapy, as well as the willingness to consume the DMB food supplement (17).

The patients were randomly assigned into three groups and received one tablet of 150 mg of DMB (standard dose), 300 mg of DMB (high dose) or a placebo for three months before each main meal. The progression of cancer was assessed by a specialized oncologist using Computer Assisted Tomography (CAT). Likewise, dietary intake, nutritional status, quality of life, physical activity and potential adverse events were monitored by specialized nutritionists and endocrinologists at the University Hospital La Paz, Madrid, Spain (17).

### 2.2 Blood Samples

Blood samples were collected in the morning (approx. 8:00 am) by trained personnel at the Hospital University La Paz Extraction Unit (Madrid, Spain) coinciding with blood tests before chemotherapy to avoid more punctures and hospital visits than necessary. Blood samples were obtained at the baseline, at mid-time and the end of the intervention in tubes containing EDTA and were centrifuged immediately at 1000 x g for 10 min. Plasma was isolated and stored at −80°C until analysis.

### 2.3 Determination of plasma cytokines and tumor cachexia factors

Based on previous reports, (21–24) relevant molecules previously described to be associated with inflammatory processes and cancer cachexia were selected for plasma analysis. Specifically, tumor necrosis factor-alpha (TNFα), interleukin (IL) 6 (IL-6), IL-1β, IL-4, IL-10, IL-15, interferon-gamma (IFN-γ), IL-15, soluble IL-6 receptor (sIL-6R), soluble TNF receptor type I (sTNFR-I) and soluble TNF receptor type II (sTNFR-II) were analyzed by the X-MAP Luminex multiplex enzimoinmmunoassay platform using specific antibodies as previously described (25). Human ciliary neurotrophic factor (CNTF) and human proteolysis-inducing factor/dermcidin (PIF) were analyzed by enzyme-linked immunosorbent assay (ELISA) following the kit instructions provided by the manufacturers. Specific bead panels and ELISA Kits used with their corresponding variation coefficients are detailed in Table 1.

**TABLE 1:** Bead panels for enzyme-linked immunosorbent assay, analytes, and variation coefficients for cytokines and cachexia tumor factors analyzed.

| **Bead Panels or Enzyme-Linked Immunosorbent Assay** | **Analytes** | **Assay CV** |
| --- | --- | --- |
| HSTCMAG-28SK-06<br>(EMD Millipore Corporation, Missouri, U.S.A.) | TNFα | 12.64 |
|  | IL-6 | 19.31 |
| | IL-1β | 7.86 |
|  | IFN-g | 7.14 |
|  | IL-4 | 16.89 |
|  | IL-10 | 8.87 |
| HCYTA-60K<br>(EMD Millipore Corporation, Missouri, U.S.A.) | IL-15 | 9.35 |
| HSCRMAG-32K-03<br>(EMD Millipore Corporation, Missouri, U.S.A.) | sIL-6R | 8.83 |
|  | sTNFR-I | 3.76 |
|  | sTNFR-II | 5.65 |
| CSB-E04527h<br>(Cusabio, Wuhan, China). | CNTF | 5.97 |
| CSB-E13626h<br>(Cusabio, Wuhan, China) | PIF | 8.54 |

Abbreviations: CNTF, Ciliary Neurotrophic Factor; IFN, Interferon; IL, Interleukin; PIF, Proteolysis Inducing Factor/Dermcidin; sIL-6R soluble IL-6 receptor; sTNFR, soluble TNF receptor; TNF, Tumor necrosis factor.

### 2.4 Statistical analysis

For each variable, the results were tested to ascertain whether they followed a normal distribution using the Shapiro-Will test. The results are expressed as the mean ± SEM. General linear mixed models of variance (GLM-ANOVA) were used to evaluate the effects of time, treatment, and time per treatment, assuming homogeneity of regression slopes. The analysis of differences in percentages was carried out through χ^2^ or Fisher’s F analysis.

To assess whether the plasma cytokine levels are independently associated with the response to the oncologic treatment, either as a reduction in cancer or a maintenance or worsening of the condition, a binary logistic regression was performed using the statistical package SPSS 25.0, with Wald backward option, and a statistical significance level of p<0.05 (IBM SPSS Inc, Chicago, IL) (26). Additionally, based on Pearson’s correlations, we examined the relationships between inflammatory variables in plasma and taste acuity, body mass index, free-fat mass, energy intake, quality of life score, as well as albumin and prealbumin in patients with cancer. Studio’s corrplot function (27) was used to express associations by correcting multiple tests with the false discovery rate (FDR) procedure (28). Plots show only significant and corrected associations. Red and purple lines indicate the correlation values, with negative correlations highlighted in red (−1) and positive correlations highlighted in purple (+1). Figure 2. All results were analyzed using the R Project for Statistical Computing (https://www.r-project.org/) (29) and a p-value < 0.05 was considered to indicate statistical significance.

## 3 Results

Firstly, we tested if plasma levels of cytokines and tumor factors were related to the success of the antineoplastic treatment, regardless of DMB consumption, by using a logistic regression analysis. We found that only levels of sTNFR-II tended to decrease with the reduction of tumor mass as measured by CAT (P=0.098).

Secondly, we analyzed changes in the circulating levels of several biomarkers of inflammation and cachexia in patients who regularly consumed DMB (standard dose and high dose) or placebo before and after three months of intervention (Table 2, Figure 1). The three groups showed a decrease in the plasma levels of IL-6, IL-1β and TNF-α throughout the intervention. The percentage change from baseline was greater in patients receiving the standard dose of DMB than in those receiving a high dose of DMB or placebo. Similar results were observed for circulating levels of the soluble receptor sIL-6R, while sTNFR-II increased only in the placebo group. In addition, the circulating levels of the tumor-derived molecule PIF decreased significantly (P=0.02) over time in the three groups, while a decrease in the CNTF concentration was observed only in the group that received the standard dose of DMB. In contrast, IL-15 increased in the DMB standard dose group but not in the other groups. The same pattern was found for the anti-inflammatory cytokine, IL-10, which increased in the groups treated with DMB but not in the placebo group.

**FIGURE 1.**
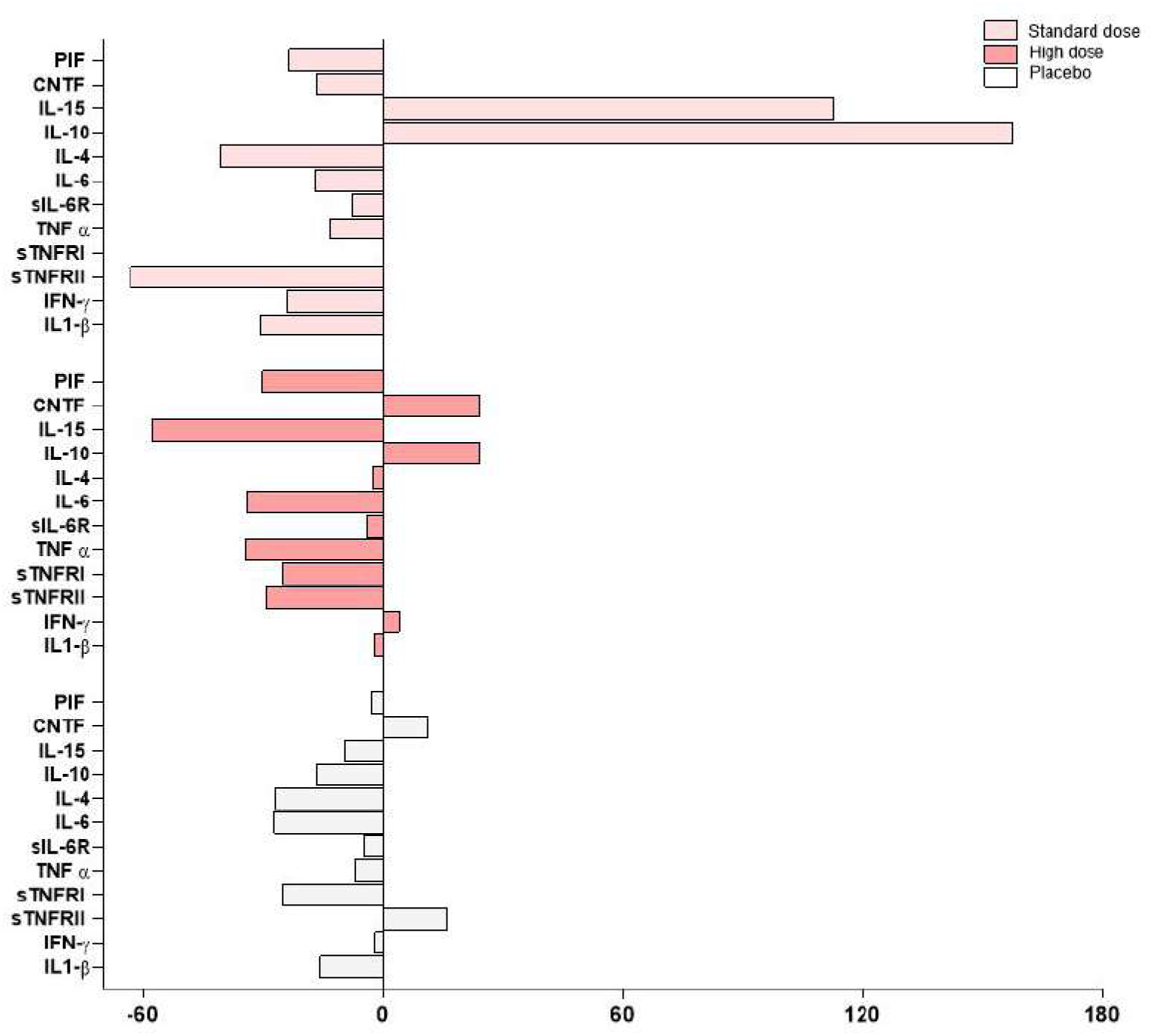
Mean change percentages of plasma levels for several biomarkers of inflammation and cachexia in malnourished cancer patients after three months of intervention with DMB or placebo. Abbreviations: CNTF, Ciliary Neurotrophic Factor; DBM, dried miracle berries; IFN, Interferon; IL, Interleukin; PIF, Proteolysis Inducing Factor/Dermcidin; sIL-6R soluble IL-6 receptor; sTNFR-soluble TNF receptor, TNF-α, Tumor necrosis factor-α.

**FIGURE 2.**
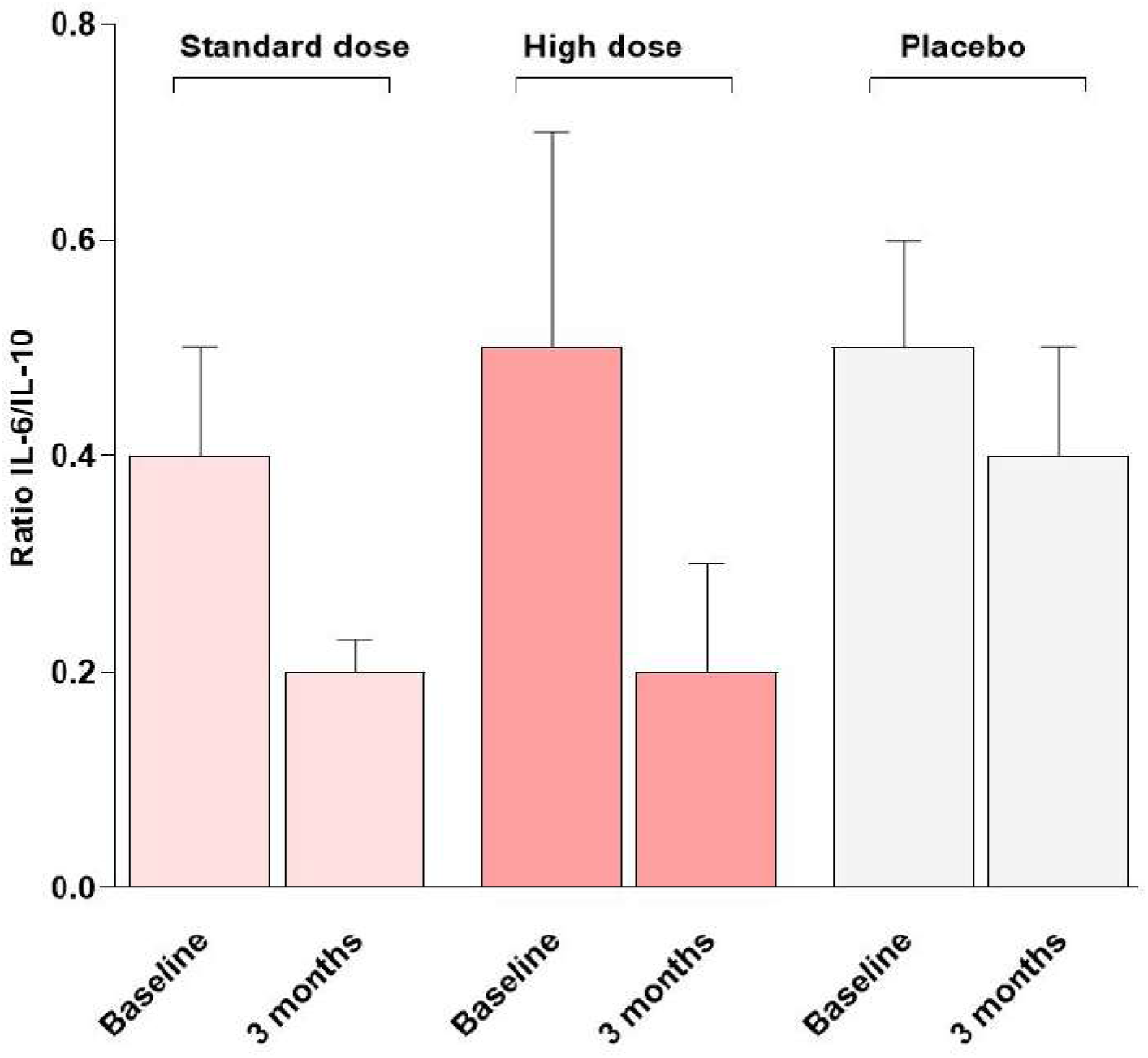
IL-6 to IL-10 ratio at the beginning of the study (baseline) and after three months of intervention with DMB or placebo.

**FIGURE 3.**
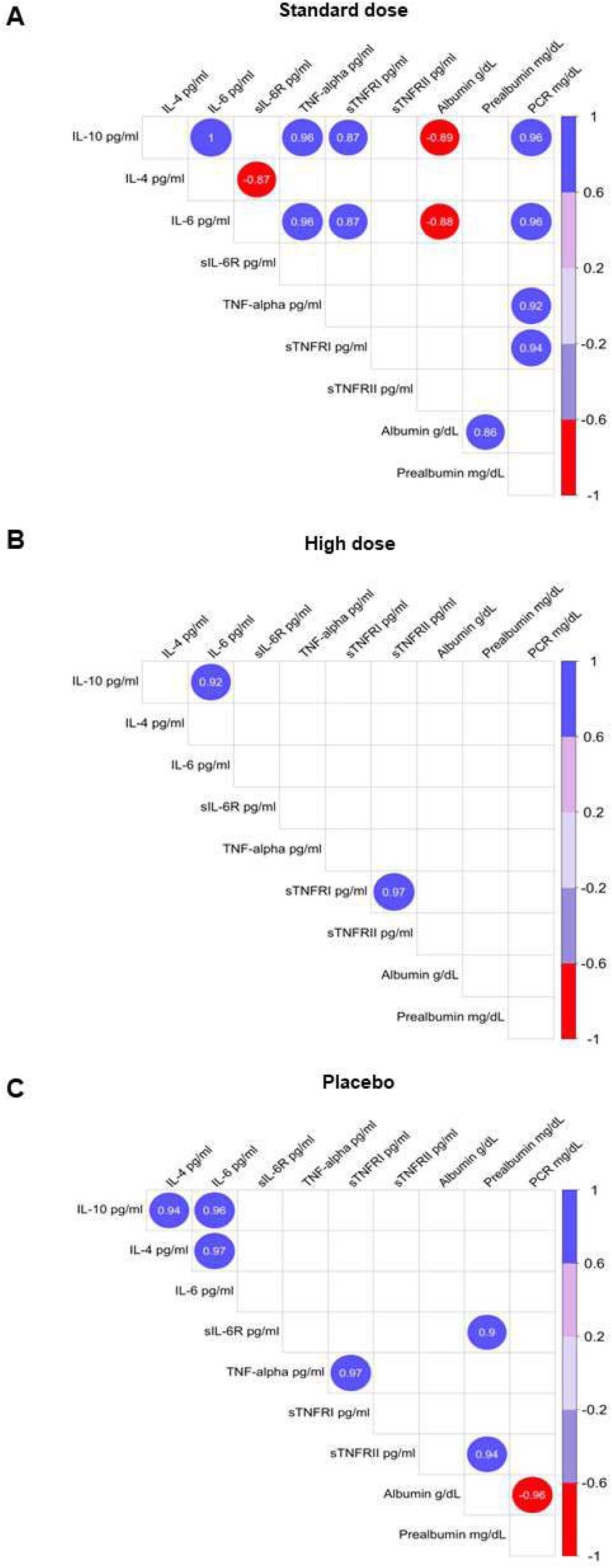
Pearson correlation coefficients between plasma inflammatory variables and nutritional status. **(A)** Standard dose of DMB; **(B)** High dose of DMB**; (C)** Placebo. Plots show only significant and corrected associations. Red and purple lines indicate the correlation values, with negative correlations highlighted in red (−1) and positive correlations highlighted in purple (+1). IL, Interleukin; PCR, protein C reactive s; TNFR-soluble TNF receptor, TNF-α, Tumor necrosis factor-α.

**TABLE 2.** Plasma levels of several biomarkers of inflammation and cachexia in malnourished cancer patients after three months of intervention with DMB or placebo.

| Variables | Standard-dose DMB<br>n=10 |  | High-dose DMB<br>n=11 |  | Placebo<br>n=10 |  | P-value |  |  |
| --- | --- | --- | --- | --- | --- | --- | --- | --- | --- |
|  | Baseline | 3 months | Baseline | 3 months | Baseline | 3 months | Time (T) | Treatment (t) | T x t |
|  | Cytokines (pg/mL) |  |  |  |  |  |  |  |  |
| IL-6 | 4.7 ± 3.6 | 3.9 ± 5.1 | 5.0 ± 2.5 | 3.3 ± 3.1 | 5.9 ± 8.8 | 4.3 ± 5.0 | 0.134 | 0.984 | 0.889 |
| IL-1 β | 6.8 ± 4.5 | 4.7 ± 2.0 | 4.3 ± 1.1 | 4.2 ± 2.9 | 4.5 ± 1.6 | 3.8 ± 0.9 | **0.093** | 0.824 | 0.501 |
| TNF-α | 18.8 ± 4.8 | 16.3 ± 9.9 | 14.2 ± 5.6 | 9.3 ± 2.8 | 14.1 ± 3.5 | 13.1 ± 4.9 | 0.297 | 0.214 | 0.652 |
| INF-γ | 39.7 ± 23.8 | 30.2 ± 15.6 | 20.0 ± 9.6 | 19.6 ± 9.1 | 18.1 ± 4.7 | 18.8 ± 10.6 | 0.226 | 0.523 | 0.209 |
| IL-15 | 11.7 ± 4.0 | 24.9 ± 33.0 | 42.1 ± 43.8 | 17.8 ± 16.6 | 10.4 ± 8.5 | 9.4 ± 2.5 | 0.205 | 0.808 | 0.389 |
| IL-4 | 46.3 ± 31.4 | 27.3 ± 12.0 | 38.2 ± 30.0 | 37.2 ± 44.0 | 71.1 ± 91.5 | 51.9 ± 66.8 | **0.04** | 0.503 | 0.374 |
| IL-10 | 10.9 ± 57.1 | 28.1 ± 48.4 | 12.8 ± 62.5 | 15.9 ± 53.1 | 17.1 ± 7.2 | 14.3 ± 16.0 | 0.47 | 0.908 | 0.554 |
| CNTF | 358.8 ± 299.7 | 300.0 ± 269.0 | 332.9 ± 258.3 | 413.3 ± 417.3 | 339.9 ± 379.3 | 378.3 ± 299.6 | 0.268 | 0.827 | 0.503 |
|  | Soluble receptors (µg/mL) |  |  |  |  |  |  |  |  |
| sIL-6R | 11.0 ± 6.3 | 10.2 ± 3.9 | 9.7 ± 3 | 9.3 ± 3.6 | 8.6 ± 3.7 | 8.2 ± 4.4 | **0.049** | 0.386 | 0.852 |
| sTNFRI | 1.3 ± 0.8 | 1.3 ± 0.8 | 1.2 ± 0.8 | 0.9 ± 0.6 | 0.9 ± 0.1 | 1.2 ± 0.5 | **0.06** | 0.377 | 0.648 |
| sTNFRII | 7.5 ± 4.8 | 4.6 ± 1.8 | 6.8 ± 3.5 | 4.8 ± 2.4 | 5.0 ± 2.0 | 5.8 ± 3.8 | **0.042** | 0.151 | 0.119 |
|  | Tumor derived factors (µg/mL) |  |  |  |  |  |  |  |  |
| PIF | 15.3 ± 7.7 | 11.7 ± 5.8 | 15.2 ± 5.2 | 10.6 ± 3.7 | 15.2 ± 5.3 | 10.2 ± 4.2 | **0.02** | 0.427 | 0.323 |

Abbreviations: CNTF, Ciliary Neurotrophic Factor; INF, Interferon; IL, Interleukin; PIF, Proteolysis Inducing Factor/Dermcidin; sIL-6R soluble IL-6 receptor; sTNFR, soluble TNF receptor; TNF, Tumor necrosis factor

Then, we determined the IL-6/ IL-10 ratio to establish the balance between pro- and anti-inflammatory cytokines. At the beginning of the study, all three groups had similar values for this parameter (0.4 ± 0.1, standard dose; 0.5 ± 0.2, high dose; 0.5 ± 0.1, placebo group). However, after 3 months of intervention, the groups that ingested DMB showed the greatest reduction in the ratio (0.2 ± 0.03, standard dose; 0.2 ± 0.1, high dose; 0.4 ± 0.1, placebo group) (Figure 2).

Finally, we tried to ascertain whether plasma cytokines (IL-1β, IL-4, IL-6, IL-10, IL-15, INF-γ and TNF-α), as well as soluble TNF-α and IL-6 receptors (sTNFR-I, sTNFR-II, sIL-6R) and C-reactive protein, were associated among them and with sensory variables, body mass index, free-fat mass, energy intake, quality of life score, as well as albumin and prealbumin. We did not find significant correlations between cytokines and sensory variables or dietary and nutritional status. However, plasma levels of C-reactive protein were positively associated with other inflammatory parameters (IL-6, TNF-α, and sTNFR-II). Additionally, IL-6 and IL-10 levels were negatively associated with plasma albumin.

## 4 Discussion

The main finding of the present study is that the regular consumption of a standard dose of DMB ameliorates several biomarkers of inflammation and cachexia in malnourished patients with cancer undergoing antineoplastic treatment and suffering taste disorders. Concerning the effects of the antineoplastic treatment, only plasma levels of the oncogene product sTNFR-II tended to decrease in patients who exhibited a reduction in tumor mass, regardless of DMB intervention. Many cancer patients, particularly those with advanced disease, experience cancer cachexia, a complex and multifactorial syndrome thought to be the result of the actions of both host and tumor-derived factors and cytokines involved in a systemic inflammatory response to the tumor (15). Cancer-associated malnutrition contributes to cachexia and can be associated with taste changes and food aversion (30–32). Early intervention with oral nutritional supplementation has been reported to be effective in reversing malnutrition, leading to a positive impact on outcomes in some patients (15). Hence, the benefits derived from the reduction in the inflammatory state observed in the present study in patients receiving the standard dose of DMB might be attributable to improved nutritional status. This improvement is due to better taste perception and increased dietary intake facilitated by DMB. Nevertheless, we were unable to detect significant correlations between plasma cytokines and sensory variables, energy intake and nutritional status, which could be due to the relatively low number of patients included in the study and the high variance of those major outcome variables evaluated. DMB is a novel food rich in miraculin, a glycoprotein that activates sweet receptors in a pH-dependent manner, as miraculin does not possess a sweet taste on its own but relies on acidification of the oral cavity to elicit a change in taste perception (33). This perception of the sour taste as sweet can persist for more than 1 hour, although the intensity of the sweet taste decreases over time (34).

Recently, we reported that regular consumption of DMB improved energy intake, fat quantity and quality, fat-free mass, and quality of life in malnourished cancer patients receiving antineoplastic treatment (16). Considering these results, a logical approach is to discern whether this improvement is reflected in the cachexia status. Indeed, improvements in cachexia parameters after nutritional interventions have already been reported (31,35). For instance, the production of proinflammatory cytokines such as IL-6, IL-1β and TNF-α is downregulated by the omega-3 PUFA eicosapentaenoic acid (EPA) in healthy people and cancer patients (31). Arachidonic acid (AA) and omega-3 PUFAs are essential for cell signaling, cell structure and membrane fluidity (36,37). Both act as eicosanoid precursors, lipid-based signaling molecules that play a key role in innate immune responses (38). AA and omega-3 PUFAs also produce a group of lipid-based pro-resolving mediators crucial for inhibiting proinflammatory signals: lipoxins, derived from arachidonic acid, and resolvins, protectins, and maresins, derived from omega-3 PUFAs. We have previously reported increased levels of selected PUFAs including linoleic acid, AA and omega-3 fatty acids (EPA and docosahexaenoic acid-DHA) following the habitual intake of a standard dose of DMB (16). These findings indicate a better food intake pattern, reflected in a better status of the PUFAs profile. Remarkably, this improvement can be attributed to supplementation with the miraculin-based food supplement DMB given that it was extended for three months, which is sufficient time for the complete renewal of the total pool of erythrocytes (39) and is in line with the amelioration observed in the biomarkers of inflammation. Furthermore, other authors reported that the effects of PIF, another cachectic biomarker, were also inhibited after EPA intake (40). Other trials aimed to identify the physiologic and clinical results of anticachexia treatment in patients with advanced cancer. MacCiò et al. treated gynecological cancer patients with megestrol acetate plus L-carnitine, a COX-2 inhibitor (celecoxib), and antioxidants versus only megestrol acetate alone; the combination treatment improved lean body mass, resting energy expenditure, fatigue, and quality of life (41).

TNF-α, IL-6 and INF-γ have been implicated as mediators of the metabolic changes associated with cancer cachexia (10,11). The present study revealed decreased levels of IL-6 and INF-γ with the time course of intervention, mainly in patients who consumed DMB. Similar results were observed for the circulating levels of the soluble receptors sIL-6R and sTNFR-II. When receptors are bound by their respective cytokines, they initiate a series of intracellular signaling cascades. Dysregulation of the expression and secretion of these cytokines and their receptors is closely linked to the pathogenesis of inflammatory diseases and cancer (42). In this sense, sIL-6R can stimulate a variety of cellular responses including proliferation, differentiation and activation of inflammatory processes, which are key in the regulation of IL-6 responses. Elevated levels of sIL-6R have been documented in numerous clinical conditions, suggesting that its production is coordinated as part of a disease response (43). On the other hand, TNF-α binds to two distinct receptors TNFR-I and TNFR-II. TNFR-I has an intracellular death domain and induces inflammation, tissue degeneration, and programmed cell death. In contrast, TNFR-II lacks a death domain and mediates primarily homeostatic effects, including cell survival, proliferation, and tissue regeneration (42). In some pathologic states, the production and release of sTNFRs may mediate the host response and determine the course and outcome of disease by interacting with TNF-α and competing with cell surface receptors (44). In the tumor microenvironment, TNF-α *via* its receptors TNFR-I and TNFR-II plays a dual role in suppressing or promoting cancer proliferation and metastasis. TNFR-I can be expressed by nearly all cells, while TNFR-II can be highly expressed by tumor cells. In malignant cells, TNFR-II promotes tumor cell proliferation and is increasingly considered an oncogene because it is overexpressed in more than 20 types of cancer (45).

Concerning anti-inflammatory cytokines, the group that received the standard dose of DMB had the greatest increase in plasma IL-10. IL-10 acts as a double-edged sword in the immune system: it is a potent anti-inflammatory and immunosuppressive cytokine but can also have immunostimulatory properties (46).

In line with these results, IL-15 levels showed a similar pattern to that of IL-10 (the greatest increase was observed in the group that received a standard dose of DMB). IL-15 can enhance the antitumor activity of the immune system. In addition, the results of clinical trials revealed that IL-15 is an effective inhibitor of tumor growth and prevents metastasis (12). Taken together, these results suggest an improvement in the inflammatory profile following the consumption of a standard dose of DMB.

The correlations observed between markers of inflammation are within the range expected in patients who suffer from a state of inflammation. Based on these data, it could be hypothesized that there is a ‘correct functioning’ of the mechanisms involved in the course of the disease and the body’s responses, which could undoubtedly facilitate therapeutic approaches. Strikingly, these correlations were observed only in the group receiving the standard dose of DMB.

Another aspect of our results that deserves attention is the positive correlation between IL-6 and IL-10 in this group. This led us to question the balance of pro- and anti-inflammatory cytokines in patients. Several studies have highlighted the importance of this balance. For example, it has been reported that the ratio of IL-6 to IL-10 is strongly correlated with injury severity and the intensity of the anti-inflammatory response after trauma (47). In addition, an imbalance in the systemic inflammatory response, marked by an increased IL-6/IL-10 ratio, is one of the possible factors contributing to the severity of primary open-angle glaucoma (48), gastric cancer (49) or COVID-19 patients (50). Understanding the interplay between IL-6 and IL-10, which reflects the balance between proinflammatory and anti-inflammatory cytokines, is critical for identifying patients in a hyperinflammatory state and will allow rational determination of the best treatment options for each patient (50).

One of the earliest events of cachexia is adipose tissue loss, often preceding skeletal muscle loss and predominantly driven by a combination of increased lipolysis and altered lipogenesis (9). Therefore, we evaluated the effect of DMB intervention on the plasma levels of CNTF, a cytokine that induces the catabolism of stored fat, skeletal muscle protein, and liver glycogen (5). CNTF and PIF are tumor-derived molecules that also induce protein degradation in skeletal muscle through the induction of the ubiquitin-proteasome proteolytic pathway (10). Remarkably, both parameters were reduced only in the DMB standard-dose group, suggesting an amelioration of cachexia in this group of patients. In contrast to the standard dose, the use of a high dose of DMB caused patients to maintain an intensive sweet taste for a long time and, therefore had a negative impact on dietary intake, which may explain why a dose greater than 150 mg is not as effective. One of the major strengths of the present pilot study is that it was carefully designed as a randomized triple-blind placebo-controlled intervention trial. In our view, although the number of patients included did not allow us to detect major differences in the evaluated parameters, it is undeniable that patients treated with the DMB supplement showed an improvement in their cachectic state in general and their immune profile in particular compared with those in the placebo group, especially at the standard dose of 150 mg (Figure 1). Consequently, this effect should not be exclusively attributed exclusively to the antineoplastic treatment alone but rather to the intervention with DMB. Thus, the success in the antineoplastic treatment was not correlated to any of the cytokines, receptors and tumor factors determined, except for sTNFR-II, which is a well-known protein highly expressed in tumor cells (51).

The major limitation of the present study is the number of subjects, although the relevance and solidity of the results are of great importance and cannot be the result of chance alone. Other studies have reported improvements in cachexia and inflammation in cancer patients after nutritional interventions, especially with long-chain omega-3 PUFAs (31,52) However, improving sensory alterations in cancer patients and promoting a better dining experience, which leads to an improvement in nutritional status and thus a decrease in biochemical parameters of inflammation and malnutrition, is a novel approach that has not been previously implemented.

This improvement also means an increase in the quality of life of patients and, as a result, a better ability to cope with harsh treatment and side effects (16).

We are aware that systemic treatment of cancer patients has a key and unique role in the amelioration of the disease. However, as all patients in our study received an antineoplastic treatment, particularly chemotherapy, the intake of DMB should be considered responsible for at least part of the observed benefits. DMB does not seem to have a direct role in tumorigenesis; however, its indirect role in improving the nutritional status and particularly increasing the essential and long-chain PUFA status could explain its effects on inflammation and cachexia. However, the dried miracle berries are rich in several bioactive compounds, mainly polyphenols, triterpenoids and amides and we cannot exclude a direct effect of DMB on cancer progression; in this sense, DMB has been shown to exhibit antitumoral activities in vitro (53,54), although studies in humans are lacking.

In conclusion, the regular consumption of a standard dose of the food supplement DMB containing miraculin along with a systemic antineoplastic treatment can contribute to improving biomarkers of inflammation and cachexia in malnourished patients with cancer and taste disorders.

## Data Availability

All data produced in the present study are available upon reasonable request to the authors

## DECLARATIONS

### Data availability statement

The data used to support the findings of this study are available from the corresponding author upon request.

### Funding information

This study is funded by Medicinal Gardens S.L. through the Center for the Development of Industrial Technology and Innovation (CDTI), “Cervera” Transfer R&D Projects. Ref. IDI-20210622. (Ministry of Science, Innovation and Universities, Spain).

### Conflict of interest statement

The authors declare no conflicts of interest.

Ethics approval and consent to participate

The study was conducted following the Declaration of Helsinki and approved by the Ethics Committee of Hospital Universitario La Paz (protocol code 6164 and Jun 23rd, 2022 date of approval).

### Informed Consent Statement

Informed consent was obtained from all subjects involved in the study.

### Clinical Trial

NCT05486260

## Acknowledgments

The authors would like to thank Medicinal Gardens S.L. (Baïa Food) for providing DMB^®^ orodispersible tablets (commercially known as TasteCare®) and for their support and technical advice.

## Notes

### Competing Interest Statement

The authors have declared no competing interest.

### Clinical Trial

NCT05486260

### Clinical Protocols

https://www.mdpi.com/2072-6643/15/21/4639

### Author Declarations

The study was conducted following the Declaration of Helsinki and approved by the Ethics Committee of Hospital Universitario La Paz (protocol code 6164 and Jun 23rd, 2022 date of approval). Informed consent was obtained from all subjects involved in the study. Clinical Trial NCT05486260

